# Cascade of care, awareness and treatment of Chronic Hepatitis B in Malaysia- findings from a community-based screening campaign

**DOI:** 10.1101/2020.06.15.20131409

**Authors:** ZZ Lim, JS Teo, AC Tan, Tan Soek Siam, Rosmawati Mohamed, KL Goh, YY Lee, WH Lai, TO Lim

## Abstract

**Introduction:** In 2016 the World Health Organization (WHO) had adopted a global strategy to eliminate Hepatitis B (HBV) by 2030 through five core interventions, the first four were preventive while the fifth is the “cascade of care”, the continuum of services that persons with chronic HBV should receive as they progress from screening to diagnosis to treatment to chronic care. We determined the prevalence of the awareness and treatment of chronic HBV in Malaysia based on a large sample data from a screening campaign.

**Methods:** A total of 10,436 subjects participated in the HBV screening campaign organized by the Hepatitis Free Pahang Malaysia (HFPM) in 2018 and 2019. All screen-positive subjects were recalled to undergo laboratory-based HBsAg and HBV DNA tests. Patients with confirmed chronic HBV were referred to local health services, while continued being monitored by HFPM.

**Results:** We estimated 13.1% of Malaysian adults aged 20 or older with chronic HBV were aware of their HBV status, and of those only 0.7% had received prior anti-viral treatment, but among those with baseline HBV DNA level>20,000 IU/ml, 15.6% were subsequently treated. Tenofovir disoproxil fumarate was the only medicine used on all treated patients. The linkage to care post-screening was broken in substantial number of patients, only 108 (54%) subjects had returned to have their HBV DNA measured and only 115 (58%) patients had subsequently sought care and were on still follow-up.

**Conclusion:** Few Malaysian adults with HBV were aware of their infection and even less received anti-viral therapy. Concerted public health efforts are urgently needed to improve HBV screening and care cascade in order to meet WHO’s targets for HBV elimination.

## Introduction

Chronic hepatitis B (HBV) is endemic in the Asia pacific region [1] including in Malaysia. A recent survey [2] estimated a prevalence of 1.2% amongst multi-ethnic Malaysian population; the disease is particularly common among Chinese with a prevalence of 2.2%. Untreated patients with HBV will progress rapidly to cirrhosis and hepatocellular carcinoma (HCC) [3]. HBV is the most common risk factor for liver cancer and accounted for more than half of all liver cancer worldwide [4]. According to the Global Burden of Disease Study 2010 [5], liver cirrhosis accounted for 42,000 years of life lost (YLLs) or 1.1% of total YLLs; it ranks ninth among the top ten major disease burden in Malaysia. Similarly, HCC is not only common (5^th^ commonest cancer), it is also the most deadly cancer in Malaysia [6], even more so than cancer of the pancreas (Table 1). Five-year patient survival with liver cancer is only 2%, this is corroborated by the only published study on HCC survival in Malaysia which reported a median survival of only 1.9 months [7]. Fortunately, therapeutic advances have rendered chronic HBV more treatable. These anti-viral therapies not only retard progression to cirrhosis, and improve survival among patients with HCC, but also cost-effective [4,8,9].

**Table 1:** Top Ten most deadly cancers in Malaysia, data from Globocan 2018[6]

| # | Cancer | Crude Incidence per 100,000 population | Crude Mortality per 100,000 population | 5 year survival probability |
| --- | --- | --- | --- | --- |
| 1. | Liver | 6.1 | 6.0 | 0.02 |
| 2. | Pancreas | 3.0 | 2.9 | 0.03 |
| 3. | Brain, nervous system | 2.4 | 2.2 | 0.08 |
| 4. | Oesophagus | 1.3 | 1.2 | 0.08 |
| 5. | Lung | 14.6 | 12.7 | 0.13 |
| 6. | Stomach | 5.0 | 4.1 | 0.18 |
| 7. | Gallbladder | 1.1 | 0.9 | 0.19 |
| 8. | Leukaemia | 5.4 | 4.3 | 0.20 |
| 9. | Ovary | 8.2 | 5.2 | 0.37 |
| 10 | Nasopharynx | 6.5 | 3.7 | 0.43 |
| 0 | All cancers | 136.8 | 82.4 | 0.40 |

In 2016 the World Health Organization adopted a global strategy to eliminate HBV and HCV by 2030 [10]. The “cascade of care”, the continuum of services that persons with chronic HBV should receive as they progress from screening to diagnosis to treatment to chronic care, is one of five core interventions to eliminate HBV. However most people with chronic HBV remain undiagnosed and untreated globally; WHO estimated only 9% of people were aware of their chronic HBV, and of those only 8% received anti-viral therapy [11].

In this study, we provide estimates of the prevalence of HBV awareness and treatment in Malaysia based on a large sample data derived from community-based screening campaigns in 2018 and 2019 organized by Hepatitis Free Pahang Malaysia (HFPM) [12]. HFPM is a Malaysia-based Non-Governmental Organization (NGO) established in 2017. The mission of HFPM is to mobilize local community to raise awareness about HBV and HCV, to provide free public screening services and to improve access to costly treatments for chronic HCV and HBV. The campaigns are entirely funded by charity and is conducted through partners in each localities or districts in Pahang and other States.

## Methods

### Study population

Study sample for this study are people who attended the HFPM’s screening campaigns in 2018 and 2019. In inviting local partners to jointly conduct these campaigns, we deliberately excluded NGOs or healthcare providers whose members or clientele potentially include persons at high risk of HBV or HCV (such as transgender, sex-worker or former prisoner/drug-user welfare groups, fisherman or other high risk trade associations, HIV/STD or opioid substitution clinics, drug rehabilitation centre or prison hospitals). The Ministry of Health’s (MOH) Medical and Research Ethics Committee approved the study and all subjects gave informed consent.

### Registration, screening and confirmatory tests

All participants of the screening campaign were required to register online (see https://hepatitisfreemsia.org.my/page.jsp?pageId=pPatientReg_Disclaimer&type=Screening). The online data system helped to support the conduct of the screening, to manage screen-positive subjects for subsequent testing and counselling, to facilitate reporting of results through short messaging service and to help capture the data for this study.

We used a low-cost point-of-care HBsAg test (POCT, AllTest Biotech Hangzhou China) to screen for HBV. The POCT have previously been validated [13]. The tests were conducted by trained nurses. The procedure was explained and verbal permission obtained from the participant prior to the testing. All screen-positive subjects were subsequently recalled to undergo testing, which were lab based serology and HBV DNA tests. Another trained nurse would counsel patients confirmed to have chronic HBV on infection transmission, risk of liver disease progression, need for monitoring and treatment. All HBsAg+ individuals were referred to the local health service for further care if they were not already receiving care. All individuals were also offered free monitoring of their HBV DNA levels by HFPM.

### Definitions

For the purpose data analysis,

- Subjects with chronic HBV were defined as those who were tested positive on the POCT and subsequently re-tested positive by lab based HBsAg test.
- Awareness of HBV status in subjects with chronic HBV was defined as answering “yes” to the nurse-administered question: “Have you ever been tested or told by a doctor that you have hepatitis B?”, and corroborated by enquiry on the circumstances leading to testing (eg pre-employment, insurance, routine health check, the name of healthcare provider providing care, if any).
- Prior treatment of chronic HBV was defined as answering “yes” to the nurse-administered question: “Were you or are you currently on anti-viral treatment prescribed by a doctor?”, and corroborated by enquiry on the name of prescribing provider, the name of the medicine(s) and on previous HBV DNA test results.
- Prospective treatment of chronic HBV was defined as patients with HBV DNA >20,000 IU/ml [14,15] measured at baseline when they were recalled to undergo confirmatory testing and who subsequently received anti-viral treatment.

### Data and Statistical analysis

The sample size for this study was based on expected chronic HBV prevalence of 2.0% and precision of the estimate as measured by its 95% exact binomial confidence intervals (CI). For a sample size of 10,000, a prevalence of 2.0% can be estimated with a 95% CI of 1.7 to 2.3, which is deemed sufficiently precise.

Participants in the screening campaign composed of a convenience sample which is not representative of the general population (female, older subjects, Chinese and Pahang residents were over-represented compare to the population). To estimate the population prevalence, post stratification [16] was used to adjust the sample totals to known population totals for age, gender and ethnicity based on the Population And Housing Census of Malaysia in 2010.

## Results

A total of 10,436 subjects participated in HBV screening campaigns in 2018 and 2019, of whom 200 were identified to have chronic HBV. Table 2 shows the characteristics of these 200 subjects. Their mean age was 52 years, with equal number of males and females, and predominantly of Chinese ethnicity (91%). Perinatal transmission is the commonest risk factor as indicated by maternal (30%) or sibling (52%) history of HBV, living with others who had HBV (55%). A high proportion of patients reported the use of acupuncture or cupping (bekam) treatment (42%), tattoo or other body piercing (25%) and needle-stick injury (38%) as possible risk factors for HBV. Drug use, transgender or sex with other men were under-reported, as expected as these are considered criminal behaviors in Malaysia and the questionnaire administration was not anonymized.

**Table 2:**
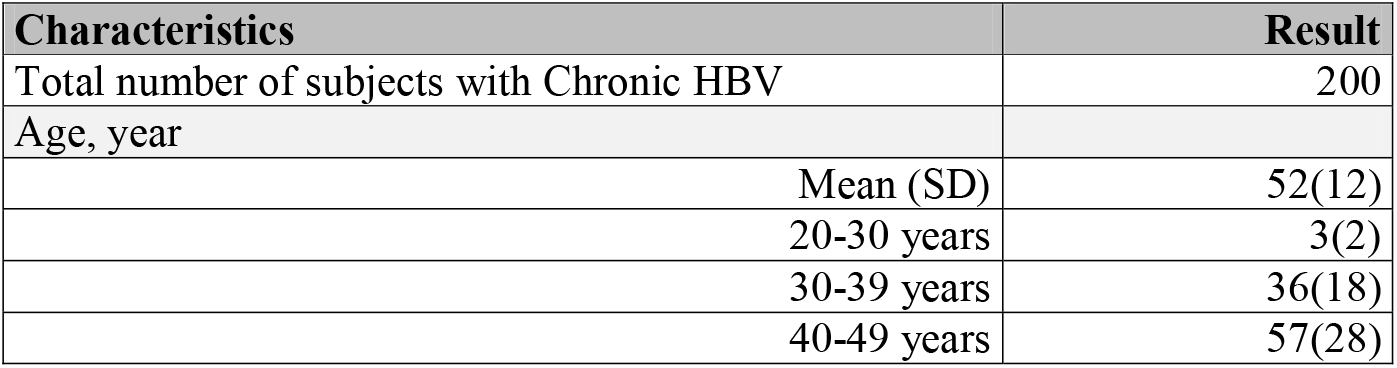
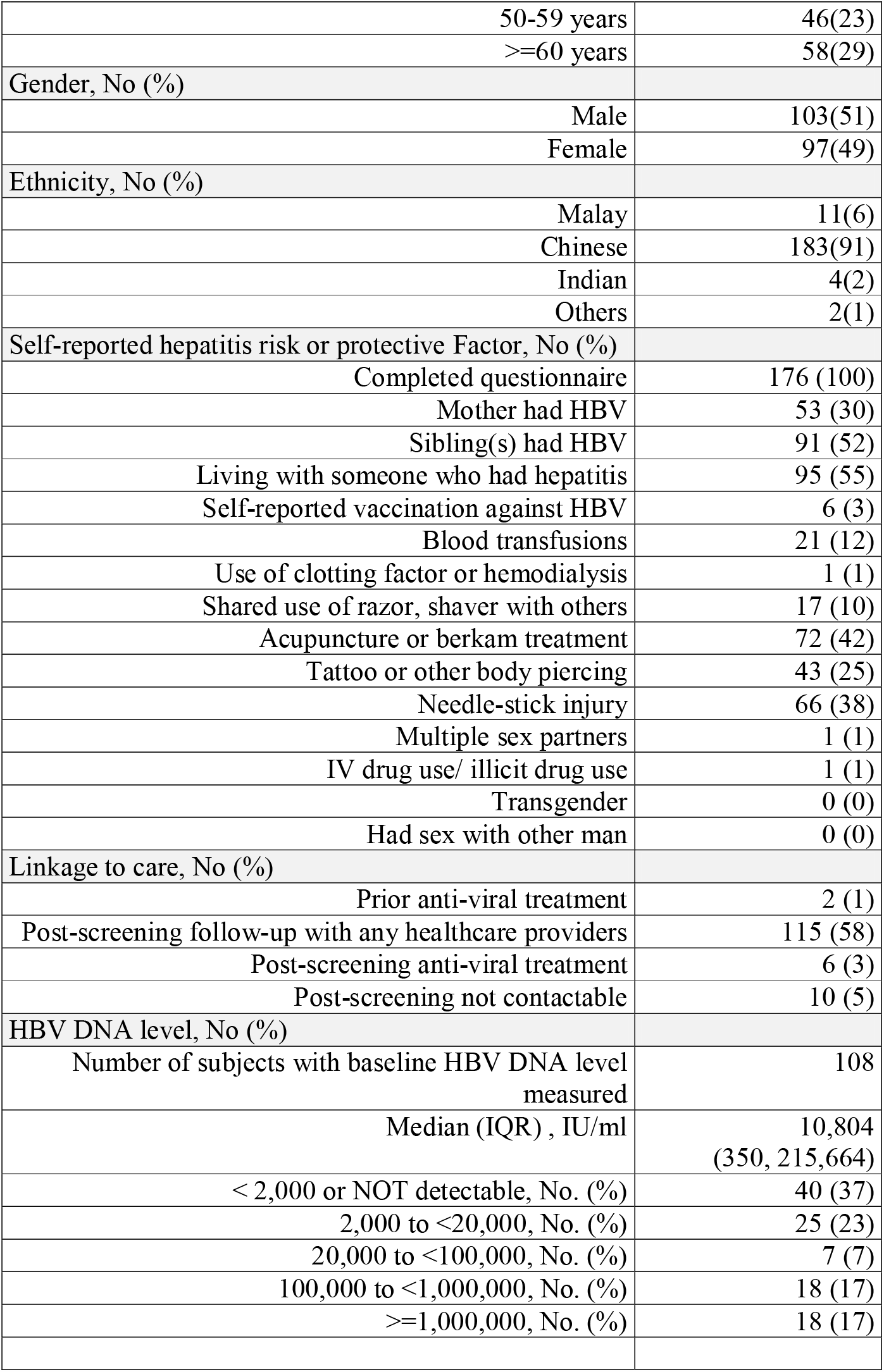
Characteristics of Chronic HBV subjects detected among 10,436 subjects screened in 2018-2019

| **Characteristics** | **Result** |
| --- | --- |
| Total number of subjects with Chronic HBV | 200 |
| Age, year |  |
| Mean (SD) | 52(12) |
| 20-30 years | 3(2) |
| 30-39 years | 36(18) |
| 40-49 years | 57(28) |

|  |  |  |
| --- | --- | --- |
|  | 50-59 years | 46(23) |
|  | >=60 years | 58(29) |
| Gender, No (%) |  |  |
|  | Male | 103(51) |
|  | Female | 97(49) |
| Ethnicity, No (%) |  |  |
|  | Malay | 11(6) |
|  | Chinese | 183(91) |
|  | Indian | 4(2) |
|  | Others | 2(1) |
| Self-reported hepatitis risk or protective Factor, No (%) |  |  |
|  | Completed questionnaire | 176 (100) |
|  | Mother had HBV | 53 (30) |
|  | Sibling(s) had HBV | 91 (52) |
|  | Living with someone who had hepatitis | 95 (55) |
|  | Self-reported vaccination against HBV | 6 (3) |
|  | Blood transfusions | 21 (12) |
|  | Use of clotting factor or hemodialysis | 1 (1) |
|  | Shared use of razor, shaver with others | 17 (10) |
|  | Acupuncture or berkam treatment | 72 (42) |
|  | Tattoo or other body piercing | 43 (25) |
|  | Needle-stick injury | 66 (38) |
|  | Multiple sex partners | 1 (1) |
|  | IV drug use/ illicit drug use | 1 (1) |
|  | Transgender | 0 (0) |
|  | Had sex with other man | 0 (0) |
| Linkage to care, No (%) |  |  |
|  | Prior anti-viral treatment | 2 (1) |
|  | Post-screening follow-up with any healthcare providers | 115 (58) |
|  | Post-screening anti-viral treatment | 6 (3) |
|  | Post-screening not contactable | 10 (5) |
| HBV DNA level, No (%) |  |  |
|  | Number of subjects with baseline HBV DNA level measured | 108 |
|  | Median (IQR) , IU/ml | 10,804<br>(350, 215,664) |
|  | < 2,000 or NOT detectable, No. (%) | 40 (37) |
|  | 2,000 to <20,000, No. (%) | 25 (23) |
|  | 20,000 to <100,000, No. (%) | 7 (7) |
|  | 100,000 to <1,000,000, No. (%) | 18 (17) |
|  | >=1,000,000, No. (%) | 18 (17) |

Of the 200 subjects with chronic HBV, only 2 (1%) had prior anti-viral treatment. Post-screening, 108 of the 200 subjects had had their HBV DNA measured. Their median HBV DNA level was 10,804 IU/ml, 37% had levels below 2,000 IU/ml while 41% had levels above 20,000 IU/ml. Only 115 (58%) individuals had subsequently sought care and were on follow-up, but only 6 (3%) received anti-viral treatment.

The prevalence of awareness and anti-viral treatment among Malaysian adults age 20 or older with chronic HBV was 13.1% and 0.7% respectively (Table 3). Of those with baseline HBV DNA>20,000 IU/ml, 15.6% had subsequently received anti-viral treatment. All were treated with Tenofovir disoproxil fumarate only.

**Table 3:**
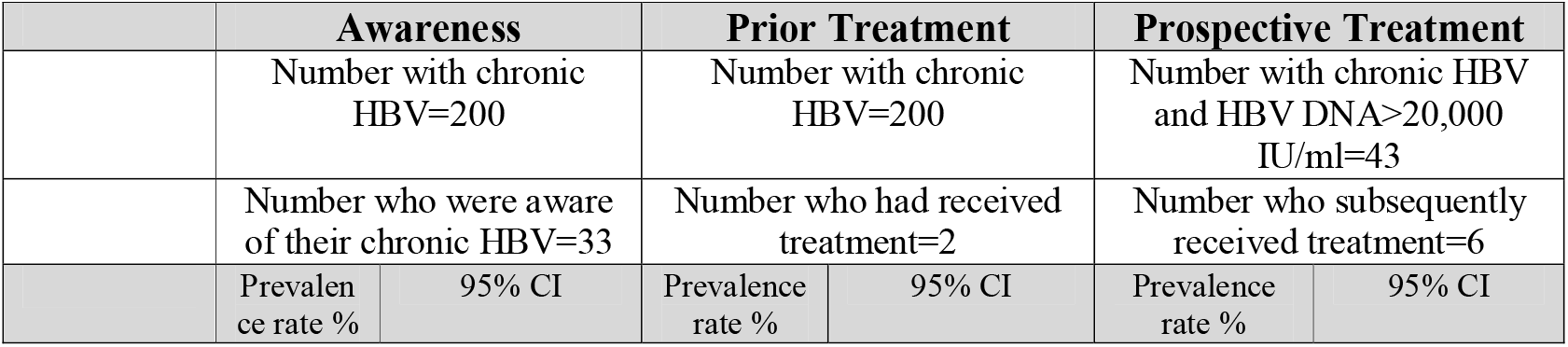
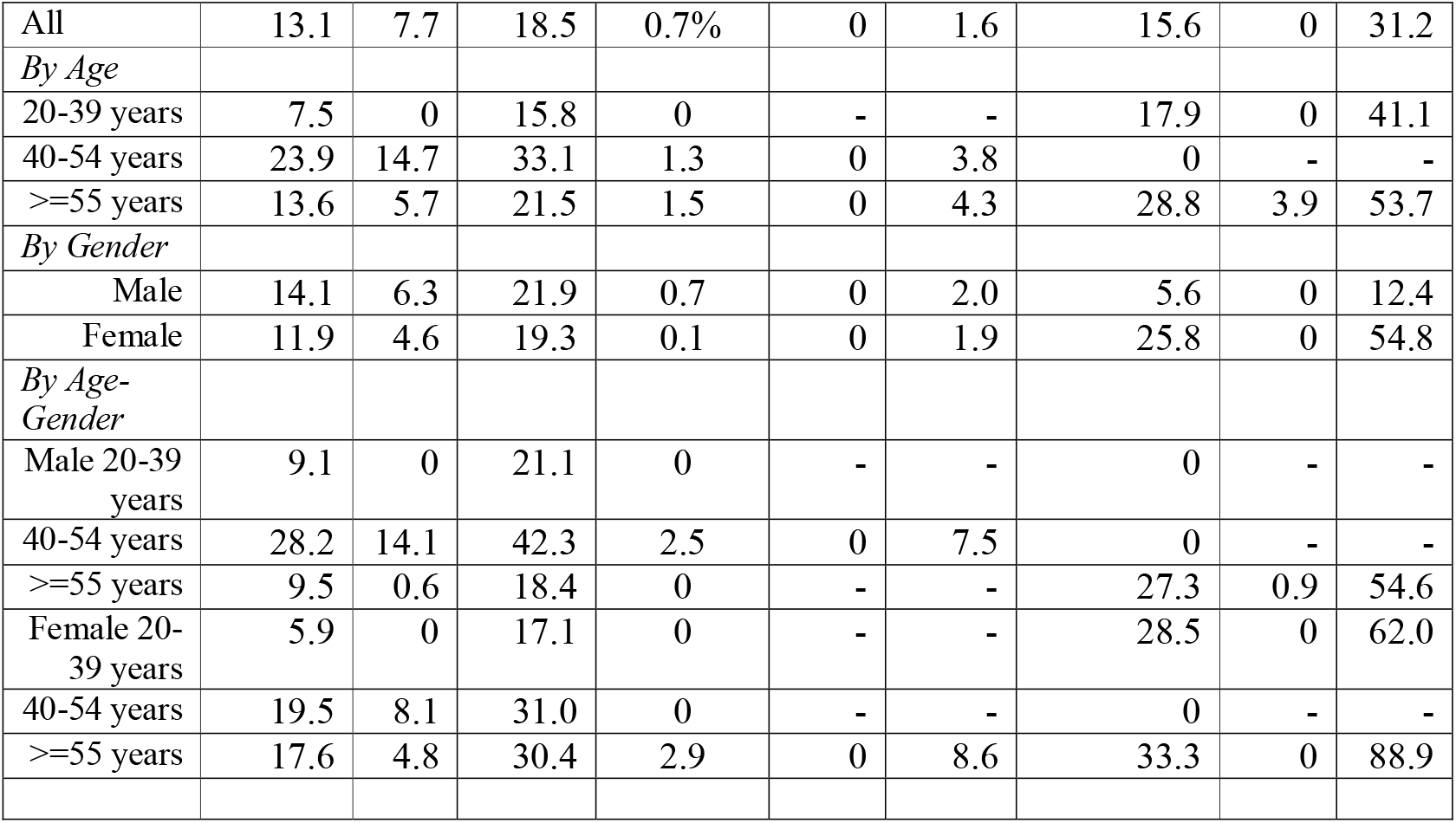
Prevalence of Awareness and Anti-viral treatment among subjects with chronic HBV in Malaysia 2018-2019

## Discussion

### HBV awareness and treatment rates

Chronic HBV is endemic in the Asia-Pacific region, though this has declined in many countries since the advent of universal vaccination of newborn [1]. However there remains a large population of adults who were born before universal vaccination were introduced in the 1990s. These adults bear the huge burden of liver cirrhosis and HCC attributable to HBV in the region; HCC ranked among the top 5 commonest cause of cancer deaths in all East and SE Asian countries without exception [17].

Nevertheless, the cascade of care, awareness and treatment of chronic HBV at the population level are surprisingly understudied in Asia. In fact, we have difficulty finding such studies beside the well-known US National Health Nutrition Examination Survey (NHANES) which has included questions on awareness of HBV only since 2013 [18,19]. Estimating HBV awareness and treatment is challenging in populations with HBV prevalence <2.5% (which is most countries in the world). The NHANES had required a rolling samples of 11,488 subjects over 4 years to identify 68 subjects with HBV of which 22 were aware and 7 treated [18]. Similarly, our survey has a sample size of 10,450 subjects to find 200 subjects with HBV, 33 were aware and 2 treated. Large scale sero-surveys have been conducted in Asia (where many countries have HBV prevalence >5%), notably in China, Korea and Mongolia [20–25]. For unknown reason, they have not reported HBV awareness and treatment rates. Sero-survey cannot determine HBV DNA guided prospective treatment rates.

The Polaris Observatory Collaborators’ study [26] have published modelled estimates of HBV awareness and treatment rates for all Asian countries in 2016. Japan, Korea and Taiwan had apparently very high awareness rates of 72%, 83% and 64% respectively. Malaysia was reported to have an awareness rate of 9% and treatment rate 5%, while in this study, the rates are 13.1% and 0.7% respectively. Despite the high percentage of self-reported intra-family HBV infection in this study population, it is concerning to find the low level of HBV awareness. We postulate that this is likely due to multiple factors in the study population and the healthcare system serving them, which warrant further study. HBV awareness and screening especially amongst women of child bearing age will help prevent mother-to-child transmission which is a vital component in HBV elimination program.

Prospective treatment rate among patients with HBV DNA>20,000 IU/ml, however is higher at 15.6% likely driven by the availability of low-cost generic Tenofovir disoproxil fumarate, which was the only anti-viral drug prescribed to all treated patients in this study. Even in the US, the awareness and prior treatment rates as determined in the NHANES survey were only 32% and 28% respectively [18].

Notwithstanding, these estimates point to the huge gap between current our health system performance and the WHO proposed targets of 90% diagnosis rate and 80% treatment rates required to eliminate HBV [11].

### Cascade of care

The low awareness, follow-up and treatment rates highlighted huge gaps in the “cascade of care” for chronic HBV. We discuss how these gaps could be closed in our healthcare setting.

- Access to HBV DNA testing to guide treatment. Only 2 screen-positive subjects had prior HBV DNA levels measured, a test which is still prohibitively costly to most patients in Malaysia. We need low-cost test, alongside a low-cost HBV treatment like generic anti-viral which drove the higher prospective treatment rate observed in this study.
- Adequately resourced and prepared local health services to deliver HBV chronic care. In Malaysia, patients with chronic HBV have hitherto been managed by specialist Hepatologists and Gastroenterologists, the supply of whom is very limited. For example, Pahang, which screened the most subjects in this campaign, has a population of 1.6 million, but only one Gastroenterologist in the public hospital and 2 in private, all are located faraway in the capital city Kuantan. This represents a bottle-neck in the cascade of care which would lead to delays in HBV follow up. In line with current WHO guideline [14], we will need to mobilize the local primary care workforce where the campaign is conducted to deliver the medical care locally for patients with uncomplicated chronic HBV.
- Non-adherence. Only 54% of screen-positive subjects who were offered free HBV DNA had come forward to be tested. And only 58% of subjects with chronic HBV had remained on follow-up with a healthcare provider despite repeated calls and reminders. Likewise, there is likely non-adherence on the part of healthcare providers [27] though this study was not designed to address this issue. Non-adherence is clearly a significant cause of failure in the linkage to care. There is room for improvement in the design and responsiveness of our health services to the needs of patients with chronic disease like HBV [28]. Similarly, we need to begin to address patients’ ignorance about HBV and the stigmatization associated with HBV, and other social and/or cultural barriers to patients seeking care [29,30,31], though again this study was not designed to address these issues.

### Study limitations

Our study has several limitations. First, the study subjects were not a probability sample and are not representative of the population. Hence, there were more female and older subjects than in the general population, and Chinese and Pahang residents were over-represented as a result of the conduct of the campaign through local NGOs, most of which were faith or ethnic based organizations in Pahang. Post-stratification was required to adjust the sampling weight to reflect the age, sex and ethnicity distribution of Malaysia. Second, subjects known to have hepatitis may be more or less willing to participate in screening. This source of bias applies to probability sample too; such subject may be more or less willing to consent to be tested in a sampling survey. The risk of this bias however is lessened in our study by pooling the data from numerous (109) screening events or venues conducted in numerous rural and urban locations spread over a wide geographical region. Third, the determination of awareness and treatment were based on self-report by patient, as is usual in all sample surveys. In our study however, instead of just accepting a Yes/No response to survey questions on awareness and treatment at face value, we had required corroborative information on the circumstances and details on names of provider/treatment and test results before accepting a “Yes” response as indicating awareness or treatment.

## Data Availability

The datasets generated during and/or analysed during the current study are available from the corresponding author on reasonable request.

https://hepatitisfreemsia.org.my/

## List of abbreviations

CI: Confidence interval
HBV: Hepatitis B virus
HCC: Hepatocellular carcinoma
HCV: Hepatitis C virus
HFPM: Hepatitis Free Pahang Malaysia
MOH: Ministry of Health Malaysia
NGO: Non-Governmental Organizations
NHANES: National Health Nutrition Examination Survey, US
POCT: Point of Care Tests
SD: Standard deviation
WHO: World Health Organization
YLL: Years of life lost

## Declarations

### Ethics approval and Consent

The Ministry of Health’s (MOH) Medical and Research Ethics Committee approved the study. All subjects who participated in the screening campaign gave written informed consent

### Consent for Publication

Not applicable. The manuscript does not report on any individual participant’s data in any form (images, videos, voice recordings etc).

### Availability of data and material

The datasets used and/or analysed during the current study are available from the corresponding author on reasonable request

### Author’s contributions

TSS, RM, GKL, LYY, LWH contributed to the subject matter expertise. They also contributed to the writing of the manuscript. LTO, LZZ, TJS, TAC conceived the idea behind this study and contributed to the study design, survey conduct, data analysis and interpretation and report writing. All authors read and approved the final manuscript.

## Acknowledgement

The authors would like to extend their sincere gratitude and appreciation to all the staff and volunteers of Hepatitis Free Pahang Malaysia for their efforts in conducting the screening campaign. We also wish to thank all those whose names are not mentioned here who render their excellent service especially during the data collection.

